# Bone2Gene: Next-generation Phenotyping of Rare Bone Diseases

**DOI:** 10.64898/2026.03.25.26349289

**Authors:** Eike Bolmer, Philipp Schmidt, Isabel Fischer, Sebastian Rassmann, Adele Ruder, Alexander Hustinx, Aron Kirchhoff, Christoph Beger, Kyra Skaf, Minu Fardipour, Tzung-Chien Hsieh, Alexandra Keller, Alberto De Rosa, Silvia Kalantari, Fabio Sirchia, Primož Kotnik, Mark Born, Benjamin D. Solomon, Rebekah L. Waikel, Tinatin Tkemaladze, Luka Abashishvili, Elene Melikidze, Anastasia Sukhiashvili, Megi Lartsuliani, Julian Nevado, Jair Tenorio, Julian Jürgens, Mona Lindschau, Christina Lampe, Shahida Moosa, Jean Tori Pantel, Larissa Mattern, Miriam Elbracht, Ho-Ming Luk, André Travessa, Julieta De Victor, Maryam Alhashim, Amal Alhashem, Noura AlKaabi, Sinem Kocagil, Emre Akbaş, Uwe Kornak, Tilman Rohrer, Roland Pfäffle, Ondřej Souček, Zdeněk Šumník, Andreas Zankl, Himanshu Goel, Markus Nöthen, Ulrike Attenberger, Amaka C. Offiah, Lothar Seefried, Hermann Josef Girschick, William Wilcox, Denny Schanze, Martin Zenker, Pablo Lapunzina, Peter Krawitz, Klaus Mohnike, Behnam Javanmardi

## Abstract

**Background:** Diagnosing the over 700 known rare bone diseases (RBDs) is inherently challenging and often requires extensive time and multiple clinical visits. Effective treatment, particularly for RBDs with approved therapies, depends on early and precise identification of the specific RBD type. Image recognition artificial intelligence (AI) has the potential to significantly enhance diagnostic processes and improve patient outcomes. Many of these disorders cause characteristic skeletal changes, especially in the hands, and are associated with growth abnormalities. Consequently, affected children routinely undergo hand radiographs for bone age assessment, making these images a widely available yet underutilized diagnostic resource.

**Materials and Methods:** We retrospectively compiled 5,623 multi-institutional hand radiographs from 2,471 patients with 45 different RBDs and 1,382 unaffected controls. We trained two deep learning models: a binary classifier to differentiate between RBD and non-RBD hand radiographs, and a multi-class classifier covering ten RBDs (or RBD groups), using 5-fold cross-validation. Preprocessing included masking, normalization, and data augmentation. Additionally, we applied occlusion sensitivity mapping to visualize class-specific features and evaluated the learned representations through cosine-based retrieval and UMAP projections of the feature space.

**Results:** The affected versus unaffected classifier achieved a balanced accuracy of 85.5% on the test dataset. The ten-class classifier reached a balanced (top-1) accuracy of 76.6%, with top-3 accuracy exceeding 90%. Disorders with highly distinctive phenotypes, such as achondroplasia, achieved accuracies above 95%, whereas phenotypically overlapping disorders, such as ACAN- and SHOX-related short stature, were more frequently confused. Feature space analysis showed that validation samples clustered closely with their respective training distributions, supporting the consistency and generalizability of the learned embeddings.

**Conclusion:** This manuscript presents a proof of principle for the development of *Bone2Gene*, a next-generation phenotyping (NGP) tool for the detection and differential diagnosis of RBDs, currently based on hand radiographs. Ongoing efforts focus on expanding the dataset to include additional RBDs or RBD groups in the current multi-class classifier for differential diagnosis and to further evaluate its generalizability. The *Bone2Gene* study is open to collaboration.

## 1 Introduction

Rare bone diseases (RBDs) comprise a heterogeneous group of disorders characterized by distinct abnormalities in bone and cartilage. According to the latest Nosology of Genetic Skeletal Disorders (Unger et al., 2023) there are currently over 700 known RBDs (i.e. around 10% of all known rare diseases) linked to mutations in more than 500 genes. The increase in the number of known disorders in the past decade reflects significant progress in identifying novel molecular causes, largely driven by advances in DNA sequencing technologies. Concurrently, the development of precision therapies has accelerated (Aartsma-Rus et al., 2022; Demirci et al., 2024). Despite these advances, many RBDs remain without curative treatments, and most patients ultimately require complex, multidisciplinary care, often including surgical interventions (Tosi et al., 2020).

Timely diagnosis is crucial for delivering appropriate care and treatment to individuals with RBDs. General practitioners, such as family physicians and pediatricians, are frequently the first point of contact; however, these clinicians often have limited experience in diagnosing, counseling, and managing rare diseases like RBDs. The growing number of conditions with known genetic causes further complicates efforts to stay abreast of the literature, even for experts. Although genetic testing is increasingly available, the diagnostic process can still take years for many patients (Hay et al., 2022), and approximately 50% of cases remain undiagnosed (Sabir et al., 2021; Scalco et al., 2026). Challenges include the interpretation of numerous variants of uncertain significance (VUS) detected through sequencing. These findings underscore that genetic testing alone is often insufficient; linking genotype (molecular data) with phenotype, often through radiologic imaging and the identification of different patterns in radiographs (Handa et al., 2023) significantly increases the diagnostic yield (Scalco et al., 2026). In fact, various international consensus guidelines recommend evaluation of skeletal abnormalities in radiographs (Savarirayan et al., 2022; Haffner et al., 2025; Dauber et al., 2026). The traditional, but still common, method to address this challenge is to work with a reference, such as the Bone Dysplasia Atlas by Spranger et al. (2018). However, this rather subjective and very time-consuming approach can lead to delayed or even wrong diagnoses.

Here, we present the first steps toward the development of *Bone2Gene*, a Next Generation Phenotyping (NGP) tool designed to differentiate characteristic radiographic patterns associated with different RBDs. Although a comprehensive skeletal survey typically includes radiographs of multiple regions, often encompassing the entire skeleton, hand radiographs (owing to their low radiation burden and routine acquisition in clinical practice) are among the most frequently obtained individual radiographic examinations in this domain (Dheeksha et al., 2023). Furthermore, recent guidelines put more emphasis on the evaluation of skeletal abnormalities in the hand and wrist radiographs to guide genetic investigation (Dauber et al., 2026). However, their potential for supporting differential diagnosis of RBDs has not been fully realized. Therefore, we chose hand radiographs as the first skeletal imaging modality for the development of *Bone2Gene*.

We aim to develop two types of deep learning models for two different stages of the diagnostic journey of RBDs. The first is a binary (affected vs. unaffected) classifier to detect dysmorphic features and identify RBD-associated hand radiographs. This model can eventually function as a screening tool at the time of radiograph acquisition. The second is a multi-class classifier to distinguish different RBDs or RBD groups. This part is similar to Face2Gene (Gurovich et al., 2019) and GestaltMatcher (Hsieh et al., 2022) for rare neuro-developmental disorders and Eye2Gene for inherited retinal diseases.

Finally, using the feature space of the multi-class disorder classifier, a higher dimensional phenotypic space was constructed to further explore phenotypic similarities and differences of different RBDs in our current cohort. This feature space (similar to that of GestaltMatcher (Hsieh et al., 2022) for faces) could eventually facilitate matching of ultra-rare cases based on hand radiographs.

Figure 1 shows a schematic overview of the proposed two-stage pipeline.

**Figure 1:**
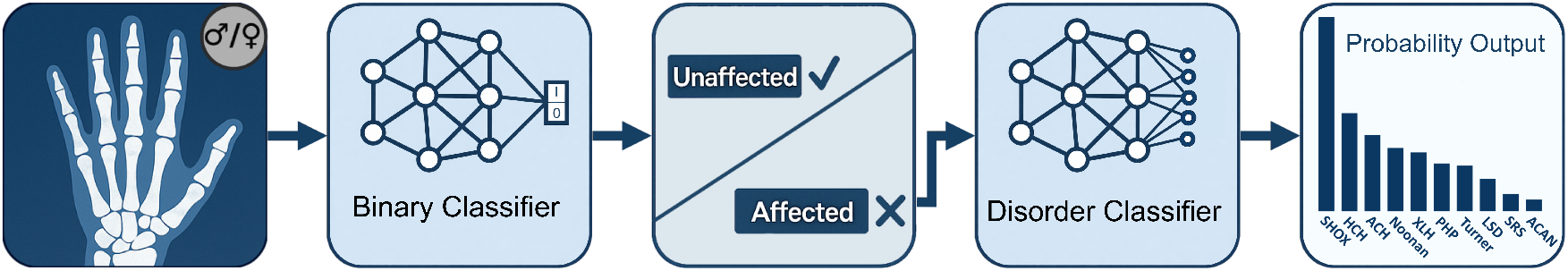
Overview of the proposed two-stage classification pipeline. Starting from a hand radiograph (with sex provided as auxiliary metadata), a binary classifier first distinguishes between unaffected and affected cases. Images classified as affected are subsequently forwarded to a disorder classifier, which assigns the sample to one of several specific disorder classes and outputs a corresponding probability distribution over the predicted diagnoses.

## 2 Materials and Methods

### 2.1 Data

The radiographic dataset used in this study was aggregated from multiple collaborating clinical centers and research partners, reflecting a broad spectrum of acquisition environments, patient demographics, and diagnostic expertise. The final dataset comprised 5,623 radiographs from 2,471 unique patients. The detailed composition of all included disorders is illustrated in Table 1. The data sources of are listed in Table A1.

**Table 1:**
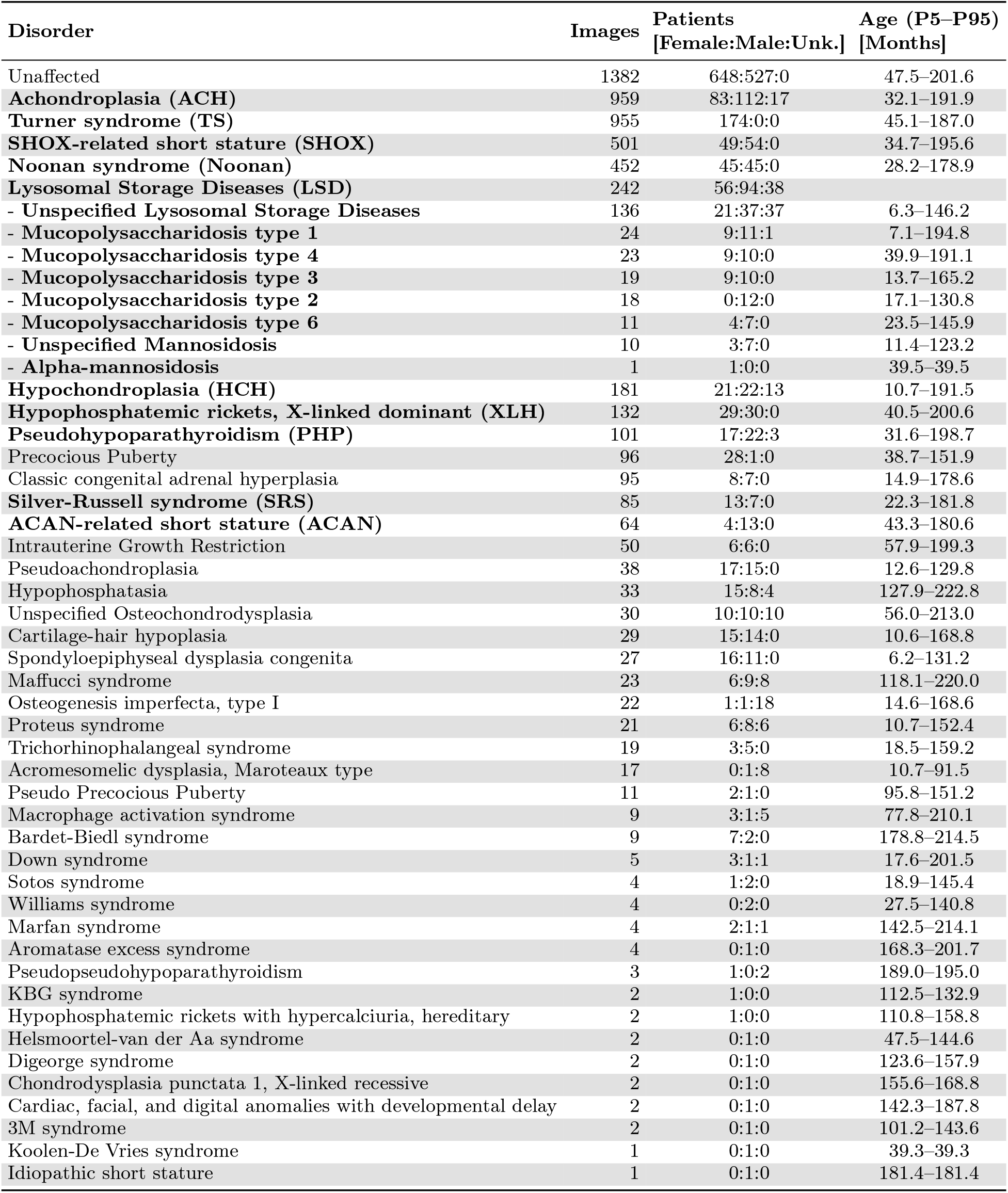
Total number of images per disorder used in this work, together with sex distribution (Female:Male:Unknown) and age range (Percentiles 5 to 95) across all images. Disorders highlighted in bold were used for our multi-class classifier. All images were used for the binary classifier.

Both male and female patients were represented in comparable proportions, and the dataset covered a wide age range from infancy to late adolescence, mirroring the diversity typically encountered in pediatric endocrine and skeletal dysplasia clinics. This diversity is essential, as many monogenic growth and skeletal disorders display age-dependent radiographic manifestations.

#### 2.1.1 Multi-class Cohort Selection

For the multi-class task, we focused on ten clinically relevant disorders with sufficient sample size. They spanned a spectrum from more distinguishable skeletal dysplasias to subtle endocrine and monogenic growth disorders:

- **Achondroplasia (ACH)** and **Hypochondroplasia (HCH)**: FGFR3-related dysplasias with characteristic radiographic patterns; Achondroplasia also serves as a sanity-check class due to its distinctive phenotype.
- **Turner syndrome (TS), SHOX-related short stature (SHOX)**, and **Noonan syndrome (Noonan)**: Common causes of short stature with overlapping but clinically important differences, where early recognition influences growth-promoting therapy and monitoring.
- **Lysosomal Storage Diseases (LSD)**: Rare but treatable conditions with dysostosis multiplex, where timely detection substantially improves outcomes.
- **Hypophosphatemic rickets, X-linked dominant (XLH)** and **Pseudohypoparathyroidism (PHP)**: Endocrine/metabolic bone diseases with characteristic hand abnormalities that guide targeted therapy.
- **Silver-Russell syndrome (SR)** and **ACAN-related short stature (ACAN)**: Subtle monogenic disorders where differentiation from other short-stature etiologies is clinically relevant.

Together, these ten disorders cover the most relevant differential diagnoses in short-stature evaluation, balancing clearly distinguishable radiographic phenotypes with disorders that benefit particularly from early identification.

### 2.2 Data Splits and Pre-processing

#### 2.2.1 Data and Splitting Strategy for the Affected vs. Unaffected Binary Classifier

For model development we derived training, validation and test sets from the multi-institutional dataset of hand radiographs described in Sec. 2.1. To avoid over-representation of any single source institution and thereby minimize source-specific bias, we implemented a sampling method that restricted the proportion of images from the largest contributors. Specifically, we iteratively reduced the share of the dominant sources by decreasing the allowance *A* of a source *s* to have a certain amount of data as affected or unaffected compared to its amount of the respective counterpart (|*N*_*s*,_−_aff_ *N*_*s*,unaff|_≤ *A*) until the maximum relative contribution 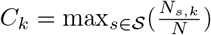 of all sources *S* and both classes *k* (aff., unaff.) is minimized across the training set, ensuring a more balanced distribution of acquisition institutions. This under-sampling strategy preserved sufficient sample size while reducing the risk that the classifier primarily learned institution-specific features rather than those relevant to the disorder.

With a resulting allowance of *A*_min_ = 285, a total of 4, 809 radiographs were available for training the binary classifier. Consequently, Datasets from sources that only consisted of affected cases were effectively sampled down to 285 images (e.g. Biomarin, that only consisted of patients with achondroplasia). The training set comprised 2, 048 images from affected individuals and 2, 761 images from unaffected controls. The validation set consisted of 499 images (206 affected, 293 unaffected), and the test set contained 378 images (248 affected, 130 unaffected). A detailed overview of all disorders contributing to the affected cohort is provided in Fig. A2.

#### 2.2.2. Data for the Multiclass Disorder Classifier

For the multi-class setting, we identified ten disorders with sufficiently large sample sizes to support a dedicated classification task. All available images for the disorder groups were included. The resulting dataset comprised 3, 649 radiographs from 936 unique patients. The most frequent disorders were ACH (959 images, 212 patients) and TS (955 images, 174 patients), followed by SHOX (478 images, 99 patients) and Noonan (452 images, 90 patients). The remaining six disorders ranged from 64 to 242 images and showed patient counts between 17 and 167. A complete overview of class frequencies and number of contributing patients is provided in Table 1.

### 2.3 Affected vs. Unaffected Classifier

To distinguish between affected and unaffected individuals, we trained a binary classification model using hand radiographs as input. The dataset was preprocessed through masking, intensity normalization, and data augmentation to improve robustness against variations from different acquisition institutions.

We implemented a binary classifier (affected vs. unaffected) with an EfficientNet b4 backbone (Tan and Le, 2019) that was initialized with weights pretrained on bone age prediction (Rassmann et al., 2024) to leverage domain-related representations. The network processed grayscale radiographs together with patient sex as an additional input feature, which was embedded through a dense layer before being concatenated with the extracted image feature from the backbone (see Fig. 2). The combined representation was passed through a series of fully connected layers with dropout regularization before the final classification layer.

**Figure 2:**
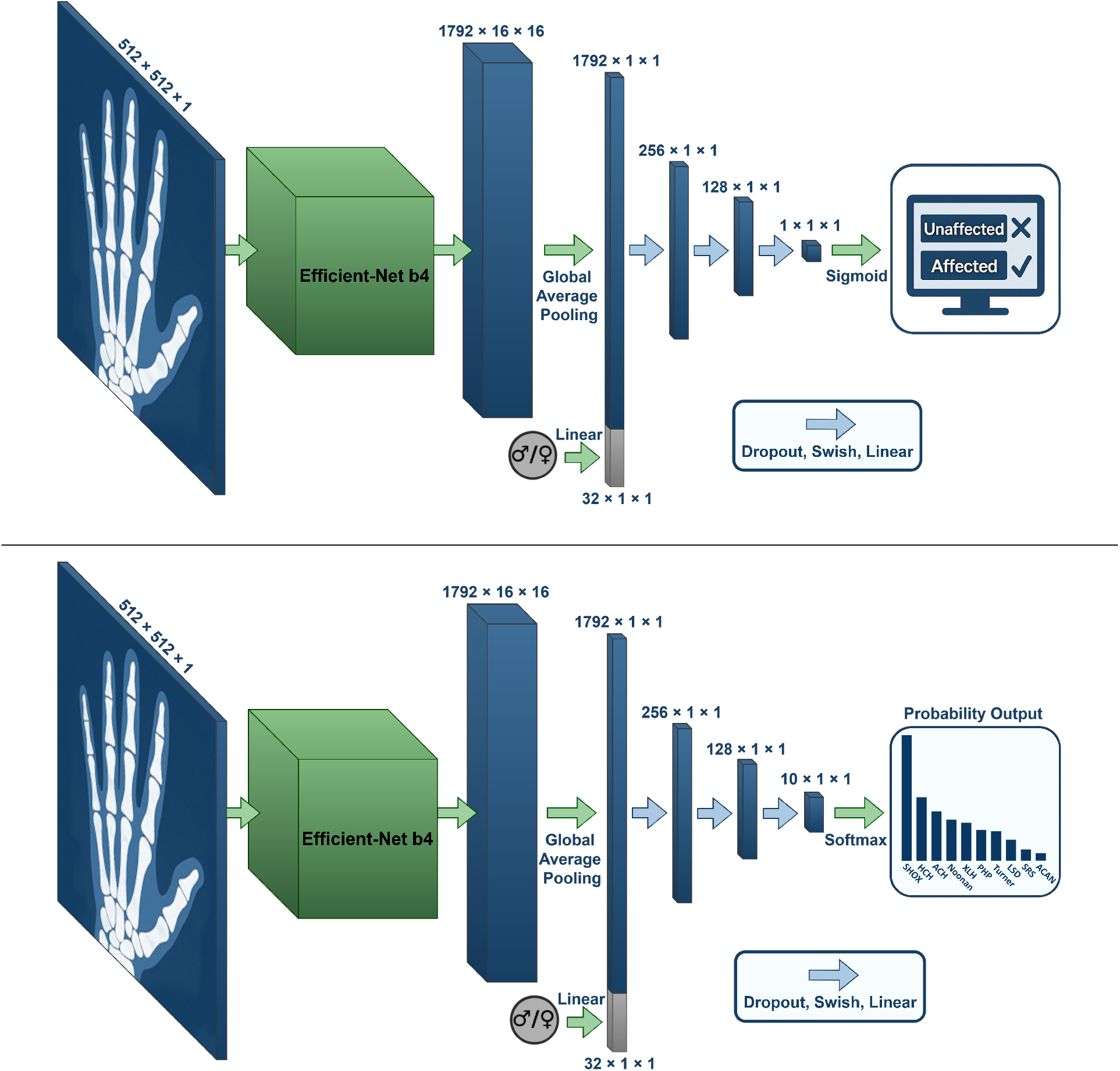
Overview of the deep learning architectures. Top: affected vs. unaffected classifier. The model processed preprocessed hand radiographs together with patient sex, extracted image features using an EfficientNet-B4 backbone, and performed binary classification to distinguish affected from unaffected individuals. Bottom: multiclass disorder classifier which followed the same architecture, but used a softmax layer with ten output classes instead of a binary head.

Training was performed with the binary cross-entropy loss and an Adam optimizer. The performance was monitored using accuracy and F1-Score on the validation set. The checkpoint achieving the highest F1-Score on the validation set was selected for testing.

### 2.4 10-Class Disorder Classifier

For disorder classification, we extended the task from a binary distinction to a ten-class setting, covering ten groups of rare bone disorders. Hyperparameters were adopted from Rassmann et al. (2024) without further tuning of the present dataset, to avoid data leakage. The preprocessing and augmentation were identical to those used for the affected vs. unaffected binary classifier, including masking, histogram equalization normalization, and geometric transformation to increase generalization.

The classifier was again based on EfficientNet-B4 with weights pretrained on bone age prediction (Rassmann et al., 2024). As in the binary model, the radiograph was processed together with patient sex, which was embedded through a small dense layer and concatenated with the image feature extracted by the backbone (see Fig. 2). The joint representation was passed through fully connected layers with dropout before the final softmax layer.

Training used categorical cross-entropy and the Adam optimizer. We performed 5-fold cross-validation. In each fold, a randomly sampled 10% of the available data was used as validation set with patient-wise splitting. The checkpoint achieving the highest F1-Score on the validation set was selected for evaluation. We note that this introduces a mild optimistic bias into the reported validation metrics.

### 2.5 Occlusion Sensitivity Mapping

To interpret the model’s decision-making and highlight image regions that most strongly contributed to the prediction, we applied occlusion sensitivity mapping (OSM), as it provides a more directly interpretable perturbation-based estimate of region importance compared to other methods used for CNN interpretability. For each disorder class, we first identified the most confidently predicted sample, defined as the image with the highest softmax probability for the corresponding class, across the five training folds of the ten-class disorder classifier.

On these images, we systematically introduced Gaussian-feathered circular occlusion masks centered at an evenly spaced grid of *n × n* equally spaced positions, covering the image uniformly without overlap. The mask had a radius of *r* = 1.2 */n* relative to the image dimensions. The procedure started with *n* = 100, which was then progressively reduced to *n* = 30 in unit steps (71 resolution levels in total), achieving finer spatial localization at lower resolutions. Resolutions below *n* = 30 were excluded, as the resulting grid became too coarse to contribute meaningfully to the composite map. At each position and scale, the occluded image was re-evaluated by the classifier, and the drop in prediction confidence relative to the unoccluded baseline was recorded. This process yielded a sensitivity map reflecting the importance of different regions of the radiograph for the classifier’s decision.

The resulting sensitivity maps across different occlusion scales were additively accumulated for each image, producing a composite OSM that emphasizes consistently influential regions. The final maps allow for visual inspection of class-specific radiographic features that drove the model’s predictions.

### 2.6 Feature Space Visualization

To assess the structure of the learned image representations, a feature vector of size 128 was extracted from the last layer before the classification layer (see Fig. 2) for each image in both the training set and the validation set. For an interpretable visualization of the feature space spanned by these vectors, we then projected these high-dimensional embeddings into a two-dimensional space using UMAP, computed on the combined set of training and validation embeddings. UMAP was applied with cosine distance on the extracted features to obtain a two-dimensional embedding, using 5 neighbors and a minimum distance of 0.8.

### 2.7 Metrics

For a given K (e.g., 1, 2, 3, 4), we determined whether the ground-truth class of the test image appeared among the top-K ranked disorder group classes. The proportion of correctly retrieved classes was first computed separately for each class and then averaged across all classes, yielding a balanced accuracy that accounts for class imbalance.

In addition to Top-*K* performance, we report standard classification metrics derived from the confusion matrix. Sensitivity (also referred to as recall or true positive rate) measured the proportion of actual positive samples that were correctly identified by the model. Specificity (true negative rate) quantified the proportion of negative samples that were correctly classified as such.

The F1-score was reported as the harmonic mean of precision and recall, thereby balancing false positives and false negatives in a single measure. As class distributions are imbalanced, class-wise F1-scores were computed and averaged to obtain a balanced estimate of overall performance.

### 3 Results

### 3.1 Affected vs. Unaffected Classifier

The binary screening model achieved an overall accuracy of 85.5%, a mean F1-score of 84.6%, a sensitivity of 82.7%, and a specificity of 90.8% on the test set. It should be noted that all disorder classes represented in the test set were also present during training. The classifier’s performance on entirely unseen disorder types cannot be directly estimated from these results and remains an open question for future evaluation. The confusion matrix (Fig. 3, 50% decision threshold) demonstrated a clear separation between affected and unaffected individuals.

**Figure 3:**
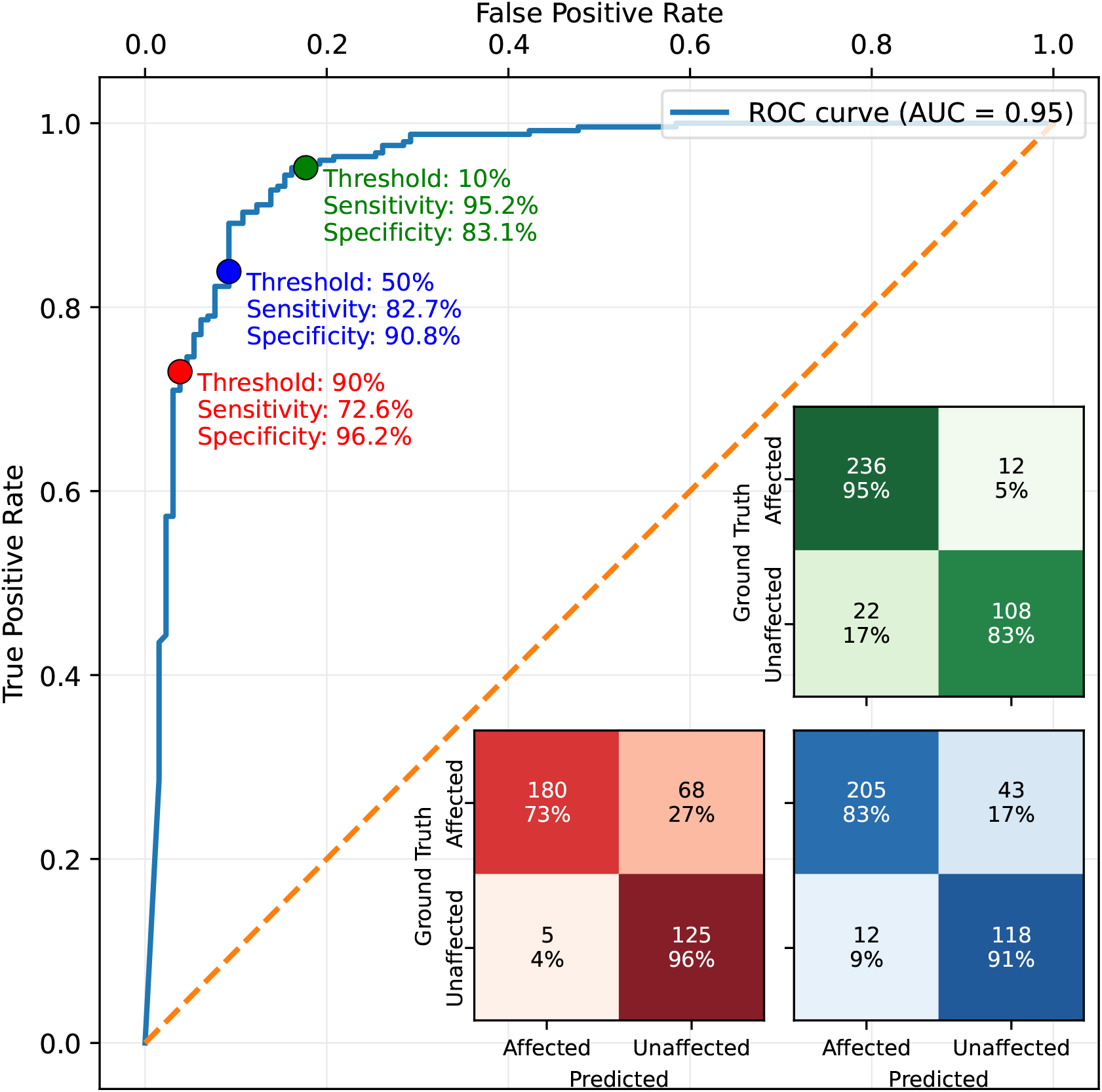
Receiver operating characteristic (ROC) curve of the affected vs. unaffected classifier. Colored markers indicate selected decision thresholds (10%, 50%, and 90%). For each threshold, the corresponding confusion matrix is shown in the lower-right corner.

**Figure 4:**
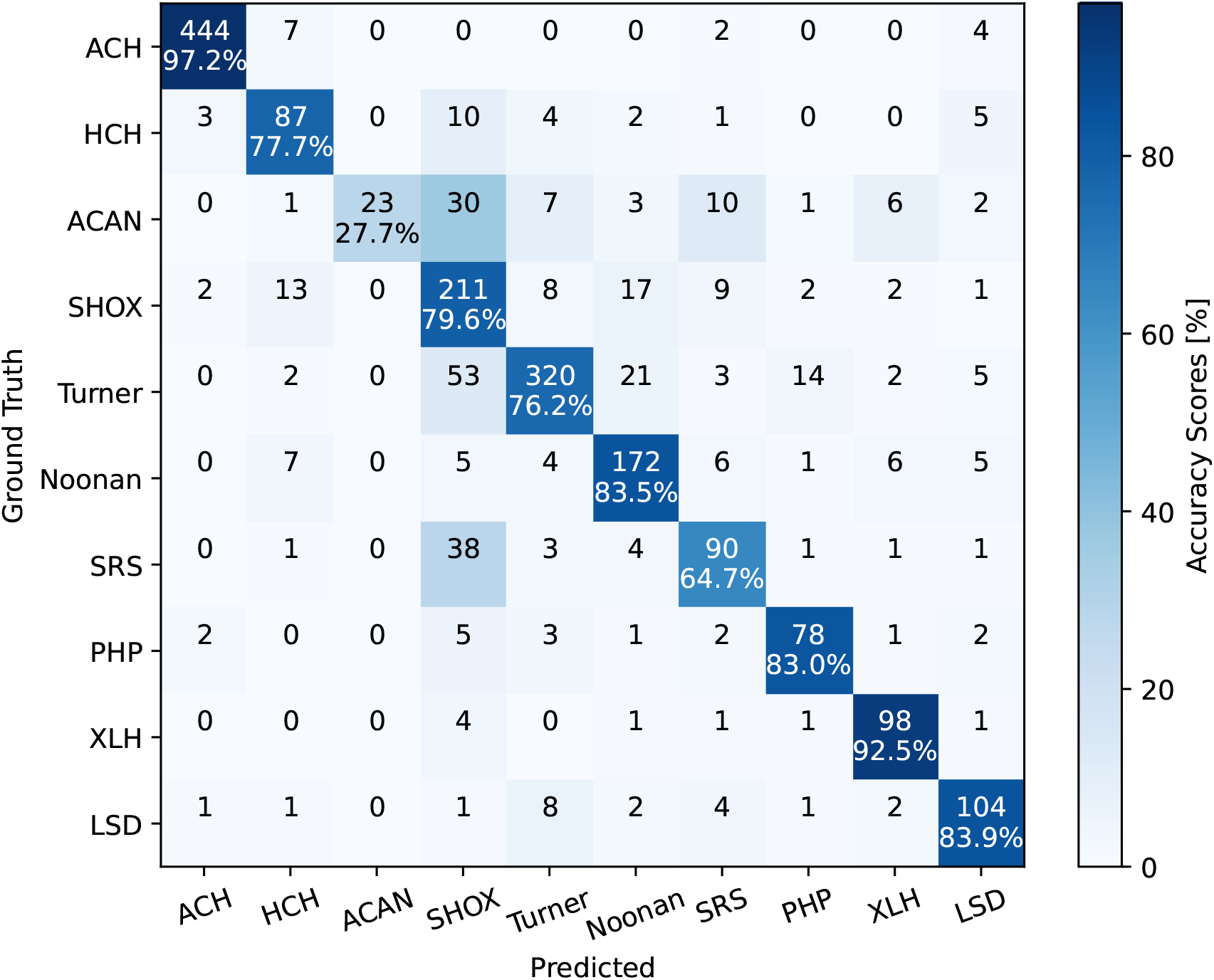
Aggregated confusion matrix of the 10-class classifier, obtained by summing predictions across all five cross-validation folds on their respective validation sets.

Different decision thresholds of 10% and 90% are also shown in the ROC curve shown in Fig. 3. With a threshold of 10%, both sensitivity and specificity increased to 95.2% and 83.1% respectively for this test set. A threshold of 90% resulted in a high specificity of 96.2%. However, sensitivity and specificity varied depending on the disorder-specific subset of the affected cohort (see Fig. A4).

### 3.2 Ten Disorder Classifier

We trained a classifier across ten rare bone disorder groups using 5-fold cross-validation. Across folds, the model reached a mean balanced accuracy of 76.59% (averaged over validation sets). In addition to Top-1 accuracy, we also evaluated Top-*K* metrics directly from the classifier output. The model achieved Top-2 and Top-3 accuracies of 86.54% and 90.36% respectively.

The per-class performance varied. Disorders with highly distinctive phenotypes, such as achondroplasia (ACH), reached accuracies above 95%. This strong performance is not surprising, given the very characteristic radiographic features of ACH and the comparatively large number of images available for this group, which makes it a reliable benchmark for the model.

Notably, hypochondroplasia (HCH) which is a phenotypically milder variant of ACH could still be classified reasonably well, reaching an accuracy of 77.7%. This is particularly noteworthy because HCH cases are frequently overlooked in routine clinical practice due to their subtler radiographic presentation (Stoll et al., 1985).

### 3.2.1 Occlusion Sensitivity Mapping

To better understand which image regions drove the model’s classification decisions, we applied occlusion sensitivity mapping. The resulting heatmaps (Fig. 5) highlighted distinct areas relevant for the model differentiating each disorder of the image from the other 9 respective RBDs.

**Figure 5:**
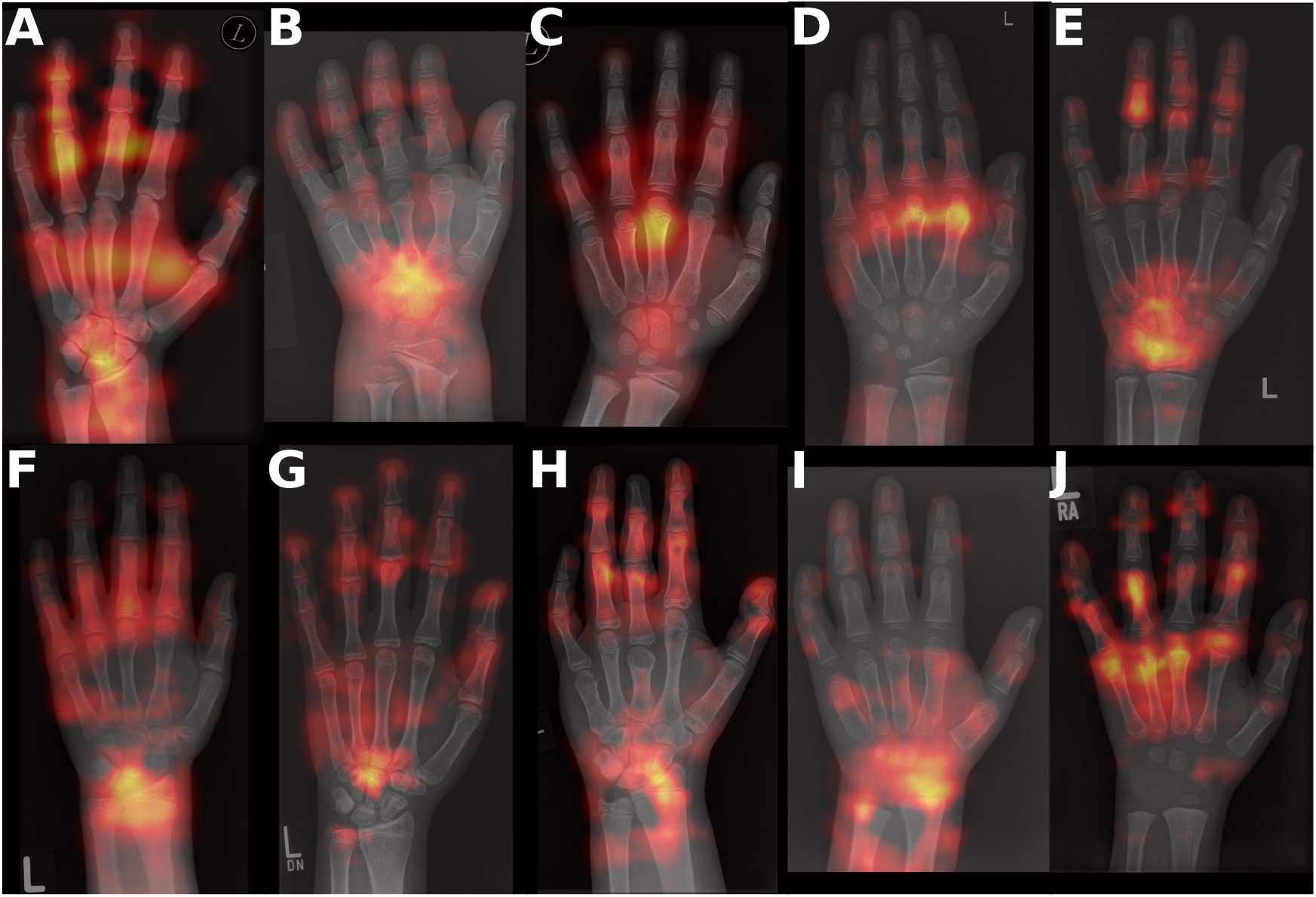
Occlusion Sensitivity Maps of sample radiographs of the ten disorder used in the disorder classifier. Highlighted (from red to bright yellow) are the areas in which the Classifier looses confidence in its classification, if that area was occluded. **A**: Hypophosphatemic rickets, X-linked dominant (XLH), **B**: Achondroplasia (ACH), **C**: Hypochondroplasia (HCH), **D**: ACAN-related short stature (ACAN), **E**: Noonan syndrome (Noonan), **F**: SHOX-related short stature (SHOX), **G**: Turner syndrome (Turner), **H**: Pseudohypoparathyroidism (PHP), **I**: Lysosomal Storage Disease (LSD), **J**: Silver-Russel syndrome (SRS).

In **XLH**, the occlusion sensitivity maps revealed predominantly diaphyseal activations. Strong relevance was observed in the diaphyses of MC2–MC5, as well as in the diaphyses of PP2–PP4 and the fourth middle phalanx (PM4). Additional sensitivity appeared in the proximal epiphysis of PD4 and the distal epiphysis of PP4. The model also responded to the metaphyses of both ulna and radius. Moreover, the carpal region showed notable activation, particularly in the area between the scaphoid, lunate, and capitate, indicating that both long-bone shafts and selected carpal structures contributed to the model’s decision-making.

In **ACH**, the heatmaps showed a broad activation pattern across the entire hand, with particularly strong signals in the carpal bones. Further relevant regions included the diaphyses of the second, third, and fifth middle phalanges (PM2, PM3, PM5) and the epiphysis of the radius. This indicated that the disorder is characterized by a more widespread pattern of relevant features rather than by highly localized hotspots.

For **HCH**, the model responses again appeared more diffusely distributed across the hand. Strong activations were seen in the distal metaphysis of MC3, but also in the diaphyses of PM2–PM5, as well as in the carpal region, particularly around the capitate and hamate.

For **ACAN**, the strongest activations appeared in the distal metaphyses of MC2–MC4 and the proximal metaphyses of PP2–PP4. Further relevant regions included the diaphyses of the ulna, radius, and proximal phalanges (PP1–PP3), suggesting that both metaphyseal and diaphyseal alterations across the radial side of the hand were central to the model’s classification.

In **Noonan**, the occlusion maps showed the strongest activations in the carpal region. Additional relevant areas included the proximal metaphyses of MC3 and MC4, as well as multiple segments of the fourth middle phalanx (PM4), specifically its diaphysis, distal metaphysis, and distal epiphysis.

For **SHOX**, the model exhibited high sensitivity in the region between the radius, ulna, and lunate, indicating that the carpal–forearm transition zone played a key role in classification. Additional strong activations appeared in the epiphyses of MC2–MC5, as well as across the entire proximal phalanges of digits 2–5, highlighting a combination of carpal, metacarpal, and proximal phalangeal features as discriminative for this disorder.

In **Turner**, the strongest activations were observed in the capitate and in the proximal epiphyses of MC2–MC4. Additional relevance appeared in the distal epiphyses of all five distal phalanges, complemented by scattered activations in other distal epiphyseal regions.

In **PHP**, the model showed widespread sensitivity across the phalanges, particularly in the distal, middle, and proximal phalanges of digits 2–4, as well as in the distal phalanx of digit 1. Additional activations appeared in the distal metaphyses of PP1 and PP5. The carpal region showed light sensitivity overall, whereas markedly stronger relevance was detected in the metaphysis and epiphysis of the radius and in the transition zone between the radius and the carpal bones. This pattern indicated that both phalangeal abnormalities and characteristic radial–carpal alterations contributed to the model’s classification.

For **LSD**, shown here in an example of a younger patient with Mucopolysaccharidosis type 2, the occlusion maps highlighted pronounced activations in the carpal region. Additional relevance appeared in the epiphyses of both ulna and radius, as well as in the transition zone between these forearm bones and the carpal bones. The diaphyses of ulna and radius also showed notable sensitivity. Across the metacarpals, light and more diffuse activations were observed as well.

Finally, in **SRS**, also illustrated with a younger patient, the occlusion maps revealed strong sensitivity in the metaphyses and epiphyses of MC2–MC5. Additional relevant regions included the proximal phalanges of digits 2, 4, and 5. This pattern suggested that characteristic alterations in the metacarpals, complemented by selected proximal phalangeal features, guided the model’s classification for this disorder.

#### 3.2.2 Feature Space Visualization

The UMAP projection (Fig. 6) of one of the five resulting cross-validation models provides a qualitative view of the learned feature space.

**Figure 6:**
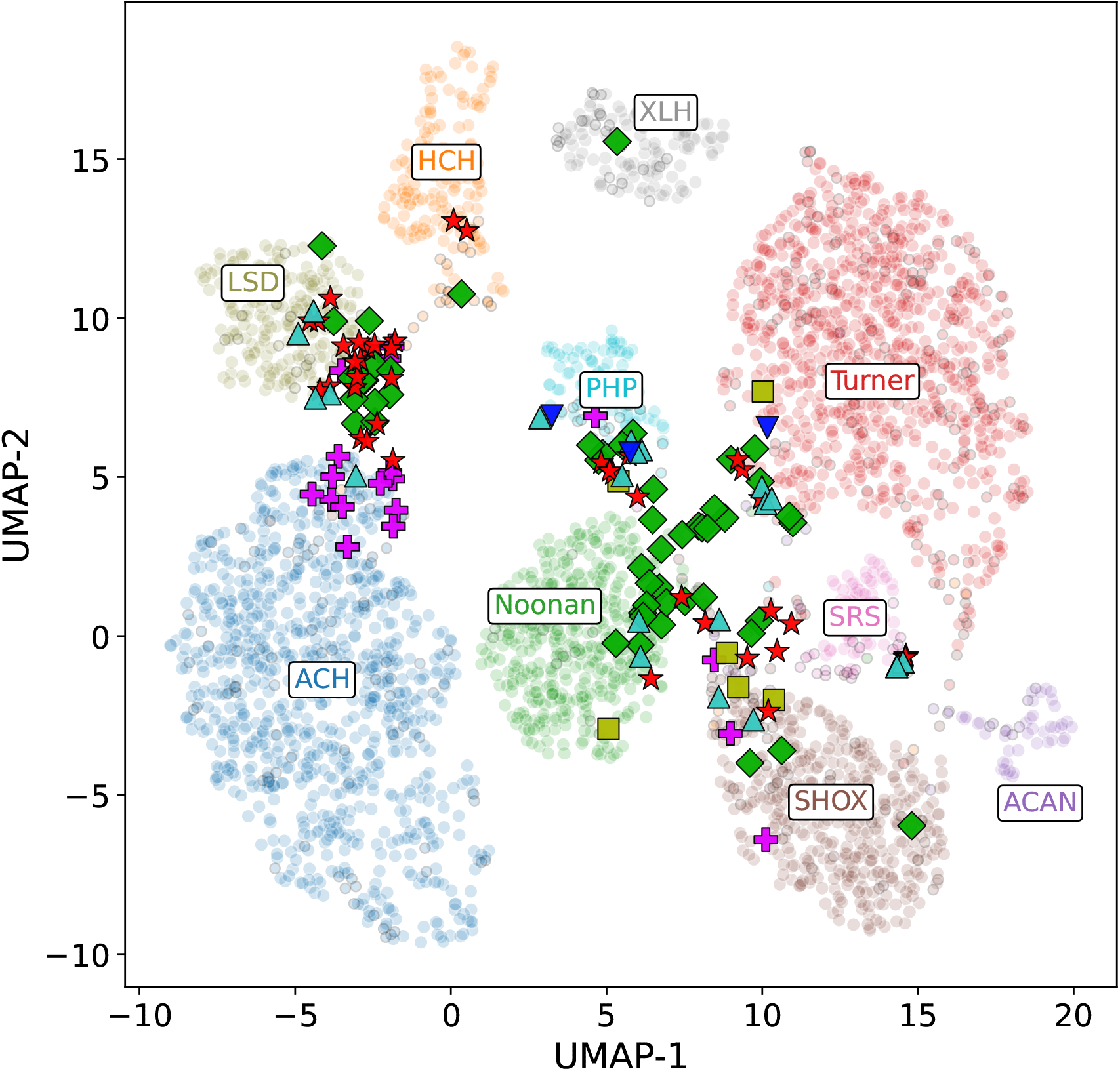
UMAP visualization of the learned feature space of the ten-disorder classifier. Training samples are shown with white borders and validation samples with black borders. In addition, samples from novel (previously unseen) disorders were projected into the same embedding space and highlighted using star-shaped markers. These include 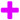 Acromesomelic dysplasia, Maroteaux type (AMDM), 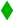 Hypophosphatasia (HPP), 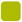 Marfan syndrome (MFS1), 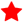 Osteogenesis imper-fecta, type I (OI1), 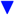 Pseudopseudohypoparathyroidism (PPHP), and 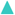 Proteus syndrome (PS).

Validation samples aligned closely with their respective training clusters, reflecting that the model captured disease-specific structure and produced consistent, generalizable representations beyond the training set.

Projection of previously unseen disorders into the same embedding space further indicated how the learned representation organized novel phenotypes relative to known classes. Notably, Acromesomelic dysplasia, Maroteaux type (AMDM) samples fell predominantly within the ACH region, which is consistent with shared growth-plate signaling involvement (e.g., NPR-related pathways). The remaining unseen disorders, including Osteogenesis imperfecta type I (OI1) and Hypophosphatasia (HPP), tended to show a preferred neighborhood (e.g., closer to LSD and Noonan syndrome, respectively), while still being more diffusely distributed across the embedding space, similar to the other novel classes. Overall, this indicated that the embedding may have captured clinically relevant similarity structure also for disorders outside the training set. At the same time, it is important to note that the model was trained with a closed-set cross-entropy objective, i.e., the representation was optimized to separate the predefined classes rather than to reserve dedicated regions for unseen disorders. Consequently, novel disorders were not expected to occupy a distinct, designated space, and their projections should be interpreted cautiously.

## 4 Discussion

The confusion matrix in Fig. 4 highlights frequent misclassifications between phenotypically similar disorders. One striking confusion is that ACAN-related short stature cases were frequently predicted as SHOX-related short stature. This may partly have been attributed to the limited number of ACAN images available in our dataset. To address this, categorical cross-entropy was computed with class-frequency-based loss weighting. On the other hand the confusion could also reflect genuine biological overlap. SHOX appears in a complex of signaling pathways influencing numerous other genes (Hoffmann et al., 2021; Marchini et al., 2016).One of its targets is the so called “SOX-trio” consisting of the transcription factors SOX5, SOX6 and SOX9 (Aza-Carmona et al., 2011; Faienza et al., 2021; Vasques et al., 2019). One of their direct targets is the activation of ACAN (Aza-Carmona et al., 2011, 2014), a gene encoding for the proteoglycan aggrecan. By building and forming the cartilage scaffold and matrix in the growth plate of long bones (Faienza et al., 2021; Kozhemyakina et al., 2015), both genes contribute to the development of a healthy bone structure and phenotypic expression. Due to the broad phenotypic ISS-spectrum of ACAN as well as SHOX (Dantas et al., 2023; Trigui et al., 2025), they might be similar in phenotypic appearance. This phenotypic spectrum can range from mild short stature to severe skeletal dysplasia with heterozygous mutations leading often to a milder phenotypic expression of ACAN and SHOX, respectively. Due to this variability and the lack of genotype-phenotype correlations it can present ambiguous phenotypes (Dantas et al., 2023; Trigui et al., 2025). There remain some open questions concerning the confusion matrix and the signalling pathways, like why there were no false-positive detections of SHOX to ACAN (0%). To find potential causalities, other signalling pathways should be considered (e.g. involving NPR2, NPPB and FGFR3). Interestingly, SHOX increases NPPB and inhibits FGFR3 expression (Vasques et al., 2019; Aza-Carmona et al., 2014).

In addition, not only ACAN-related cases but also cases from other disorder groups were occasionally misclassified as SHOX. The two false-positive predictions with the highest model confidence for ACAN, HCH, SRS, and Turner are illustrated in Fig. 7.

**Figure 7:**
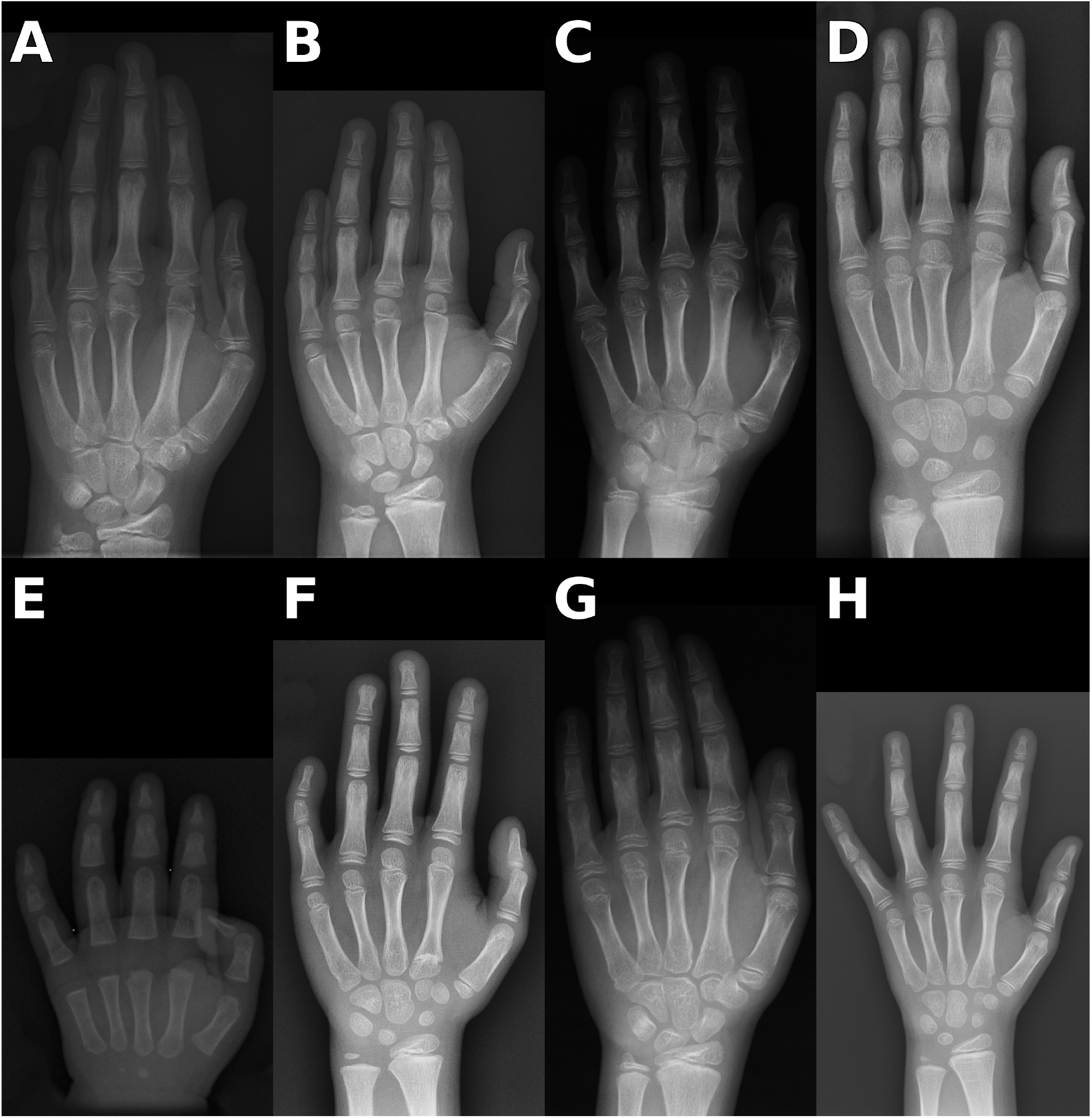
Images that were predicted as SHOX-related short stature, but were in fact cases of ACAN-related short stature (ACAN) (**A, B**), Hypochondroplasia (HCH) (**C**,**D**), Silver-Russel syndrome (SRS) (**E, F**), Turner syndrome (Turner) (**G, H**).

## 5 Conclusion

In this work, we presented the first steps toward the development of *Bone2Gene*, a deep-learning framework for detection and differential diagnosis of rare bone diseases (RBDs) based on hand radiographs. As a first step towards a clinically applicable decision-support system, we developed a binary classifier that distinguished between RBD and non-RBD radiographs with a balanced accuracy of 85.5%. Building on this, we trained a ten-class disorder classifier that reached a balanced accuracy of 76.6% and Top-3 accuracies above 90% across ten clinically relevant groups. Disorders with highly distinctive radiographic phenotypes, such as achondroplasia, were recognized with accuracies above 95%, whereas phenotypically overlapping entities, for example ACAN- and SHOX-related short stature, were more frequently confused.

Beyond overall performance metrics, we systematically analyzed the learned feature space and model behavior. Occlusion sensitivity mapping further revealed disorder-specific patterns of relevance across metacarpals, phalanges, carpal bones, and the distal forearm. These maps highlighted that the models did not rely on a single discriminative “spot,” but instead captured distributed phenotypic signatures characteristic of individual RBDs.

Taken together, our results demonstrate that AI-based screening and classification tools such as *Bone2Gene* can capture disease-specific radiographic features and may support earlier recognition of RBD patients.

## 6 Future Work

Future work will focus on several directions. One continuous effort is to expand the algorithm to more RBDs, i.e. to extend the multi-class classifier beyond the ten disorders presented in this manuscript. That requires continuous data collection. On the technical side, we aim to improve the feature space by incorporating contrastive learning, which may enhance retrieval performance and facilitate the positioning of previously unseen or ultra-rare disorders within the learned embedding space. Also, we will aim at extending our algorithms to learn features from additional skeletal regions other than hand and wrist.

Finally, before deployment in clinical practice, *Bone2Gene* will require prospective validation in independent cohorts, careful assessment of robustness across institutions, and user-centered evaluations. Addressing these aspects, together with transparent model explanations and appropriate regulatory and ethical frameworks, will be essential to translate NGP tools from proof-of-concept to reliable clinical decision support systems.

## Ethics

The Bone2Gene study has been approved by the ethics commission of the University Hospital Bonn on 09.09.2021 (Ref. No. 386/17). Informed consent was not required because only de-identified radiological images were analyzed. The study was conducted in accordance with the Declaration of Helsinki.

## Competing interests

B.J. S.R, P.K., and K.M. are co-inventors of a pending patent application containing parts of the methods described in this manuscript.

## Funding Statement

The Bone2Gene project (PI: B.J., https://bone2gene.org) is funded by the German Federal Ministry of Research, Technology, and Space within the GO-Bio initial program (https://www.go-bio.de/gobio/de/gefoerderte-projekte/gobio-initial/_documents/bone2gene.html).

## Acknowledgment

We thank BioMarin Pharmaceutical for supporting this study by providing imaging data.

## Data Availability Statement

The dataset used in this study is currently not openly available.

## Appendix

**Table A1:** Number of images per data source used in this study.

| Data Source | Number of Images |
| --- | --- |
| University Hospital Magdeburg | 2341 |
| BioMarin | 796 |
| University Hospital La Paz, Madrid | 785 |
| Kinderzentrum Am Johannisplatz, Leipzig | 512 |
| Los Angeles Children’s Hospital (Gertych et al., 2007) | 285 |
| Radiological Society of North America (Halabi et al., 2019) | 285 |
| University Hospital of Bonn | 192 |
| Publications | 188 |
| University of Pavia | 144 |
| Ljubljana University Medical Centre | 50 |
| University Medical Center Hamburg-Eppendorf | 28 |
| University Hospital of Giessen | 17 |

**Figure A1:**
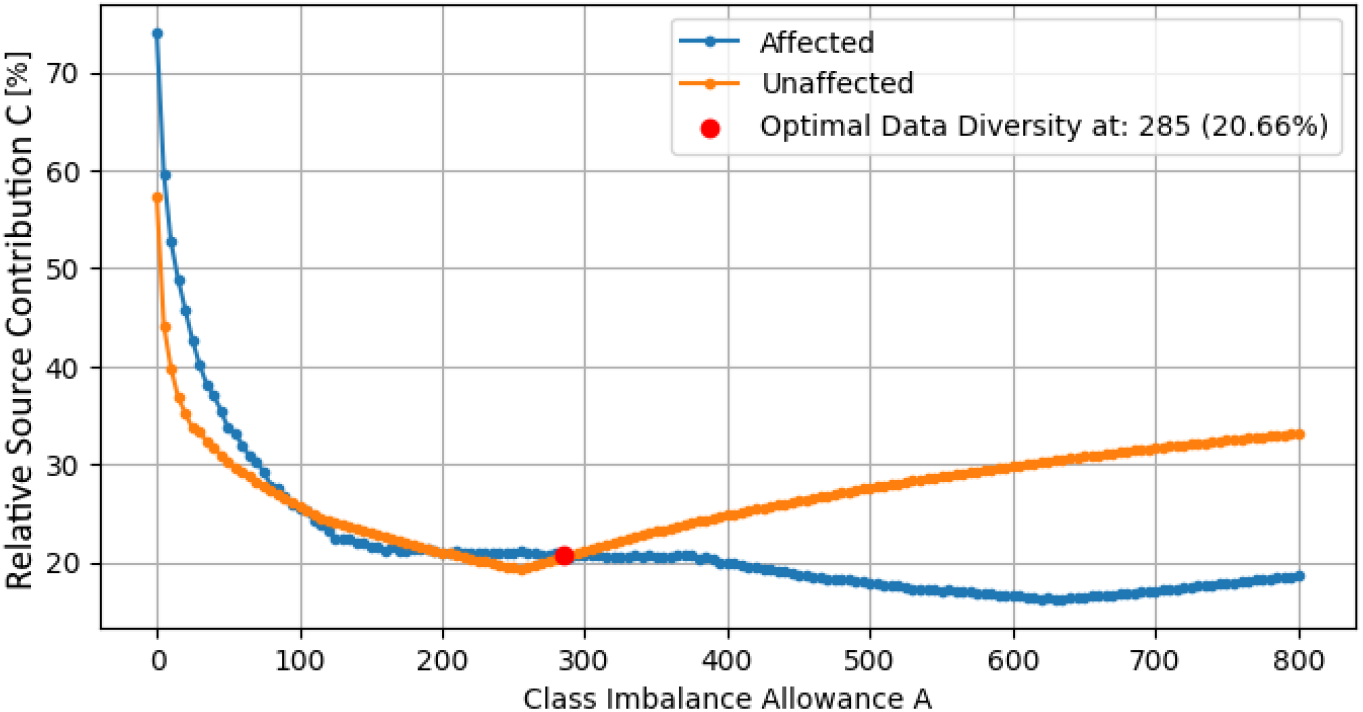
Different Class imbalance allowances *A* result in different Data Set diversities. The highest diversity or lowest share of the largest data set class imbalance from one institution of *C* = 19.79% is achieved with a class imbalance allowance *A*_min_ = 285.

**Figure A2:**
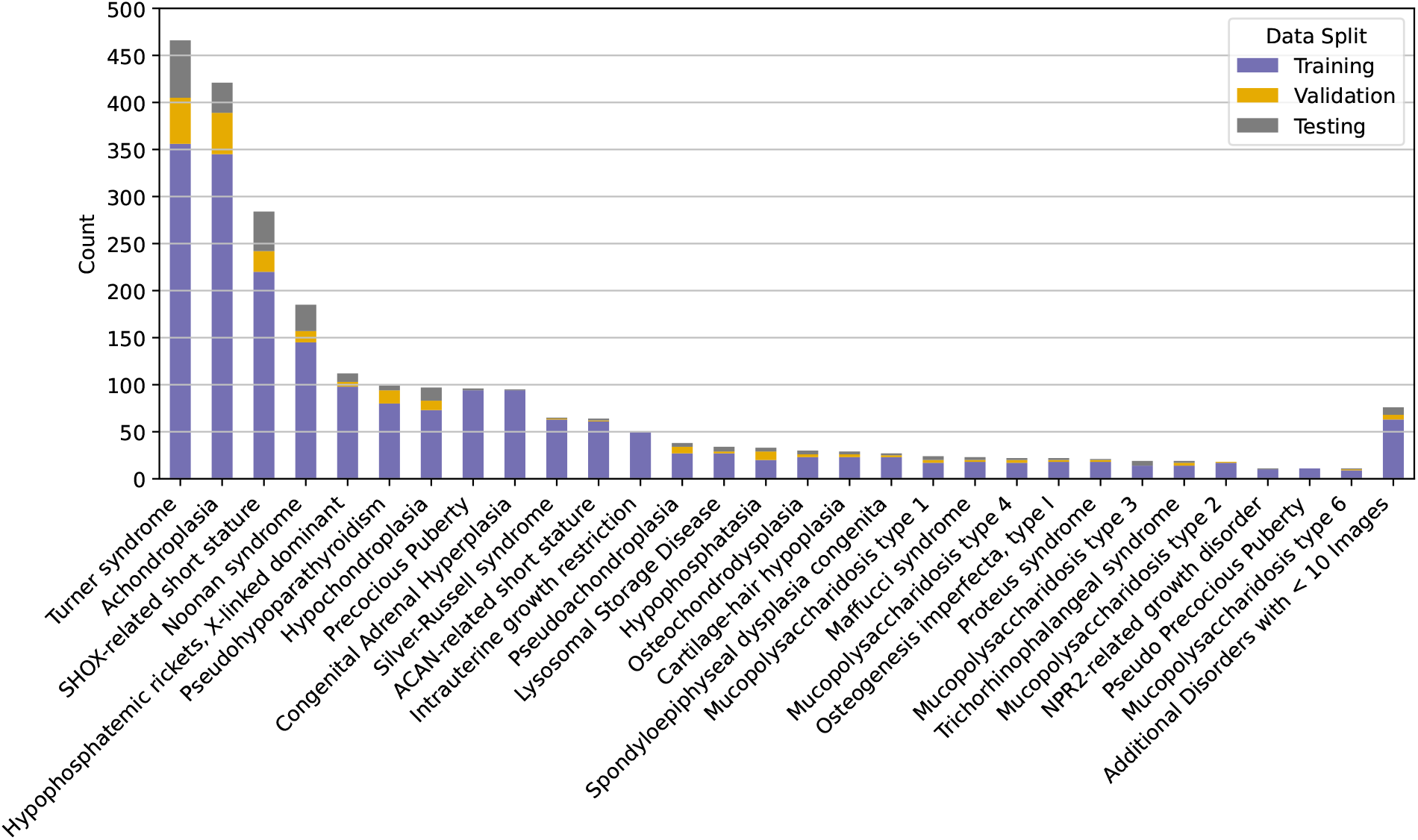
Image counts of hand radiographs of different disorders used for the binary classifier in addition to 2,761 images from unaffected controls.

**Figure A3:**
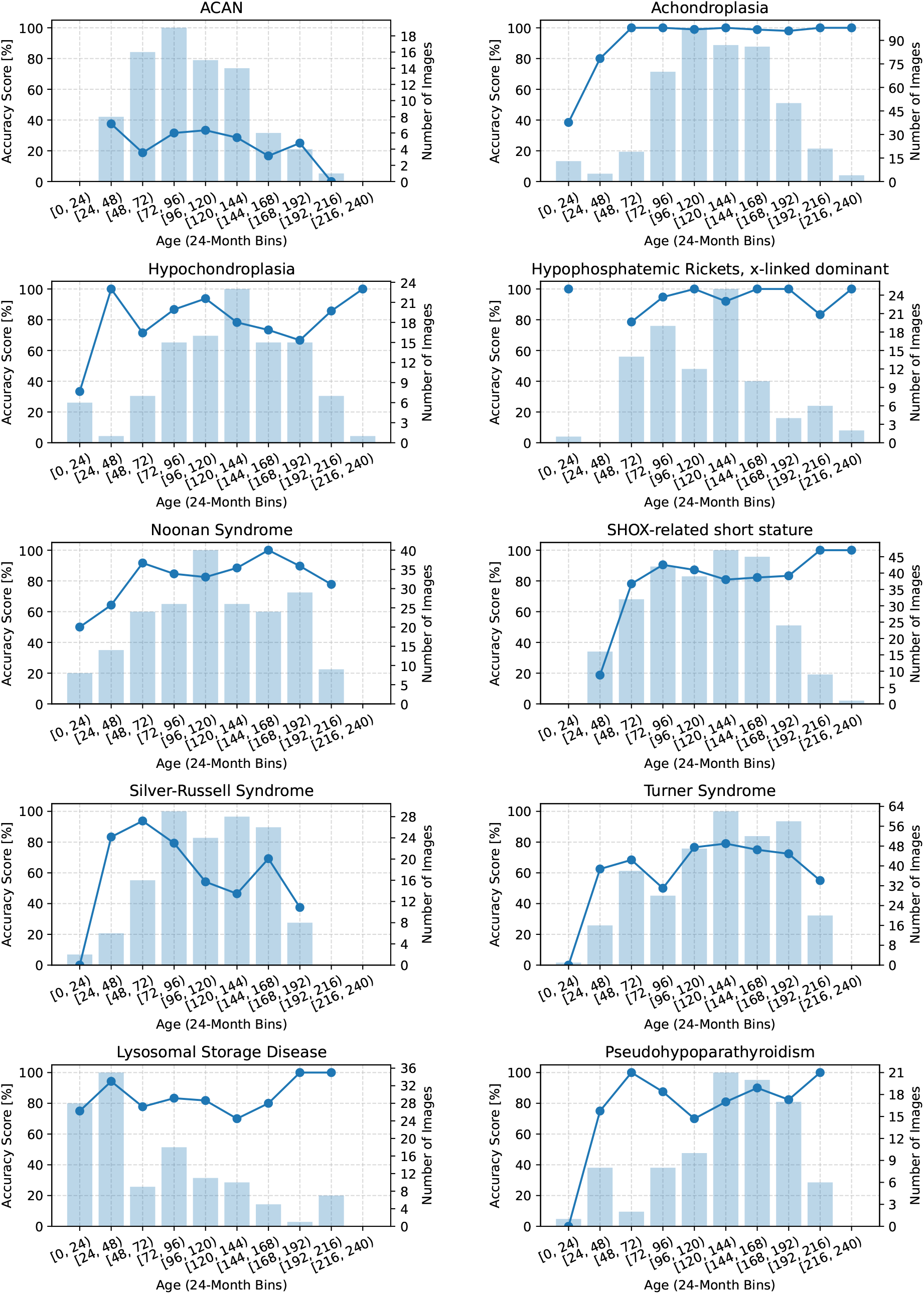
Recall by ages (24-month bins) and number of images per bin for all disorder classes.

**Figure A4:**
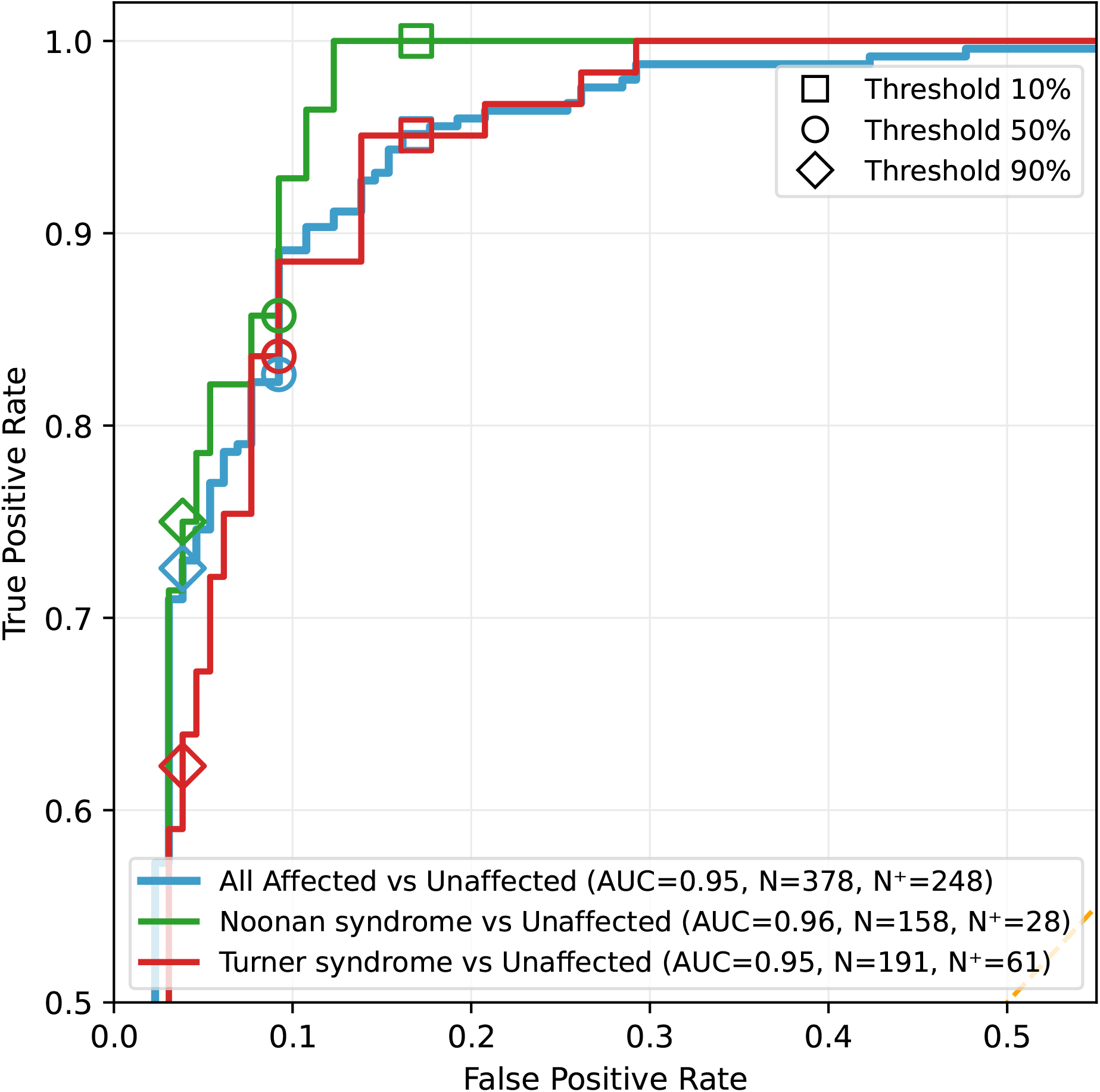
Zoomed in Receiver operating characteristic (ROC) curve of the affected vs. unaffected classifier. Colored lines indicate selected subsets of the affected cohort (blue): Turner syndrome (red), Noonan syndrome (green). Different decision thresholds are marked for each disorder.

## Notes

### Funding Statement

The Bone2Gene project (PI: B.J., www.bone2gene.org) is funded by the German Federal Ministry of Research, Technology, and Space within the GO-Bio initial program (https://www.go-bio.de/gobio/de/gefoerderte-projekte/gobio-initial/_documents/bone2gene.html).

### Author Declarations

The current study is part of the Bone2Gene project which has been approved by the ethics commission of the University Hospital Bonn on 09.09.2021 (Ref. No. 386/17). Informed consent was not required because only de-identified radiological images were analyzed. The study was conducted in accordance with the Declaration of Helsinki.

